# Measuring the Impact of Medical Cannabis Law Adoption on Employer-sponsored Health Insurance Costs: A Difference-in-Difference analysis, 2003-2022

**DOI:** 10.1101/2024.04.26.24306383

**Authors:** Mitchell L. Doucette, Dipak Hemraj, Emily Fisher, D. Luke Macfarlan

## Abstract

**Introduction:** Recent studies suggest that medical cannabis laws might contribute to a reduction in health insurance costs within the individual health insurance markets at the state level. We investigated the effects of adopting a medical cannabis law on the cost of employer-sponsored health insurance.

**Methods:** We analyzed state-level data from the Medical Expenditure Panel Survey - Insurance Component (MEPS-IC) Private Sector spanning from 2003 to 2022. The outcomes included log transformed average total premiums per employee for single, employee-plus-one, and family coverage plans. We utilized the Sun and Abraham (2021) difference-in-difference (DiD) method, looking at the overall DiD and event-study DiD. Models were adjusted for various state-level demographics and dichotomous policy variables including whether a state later adopted recreational cannabis as well as time and unit fixed effects and population weights.

**Results:** For states that adopted a medical cannabis law, there was a significant decrease in the log average total premium per employee for single (-0.034, standard error [SE] = 0.009) and employee plus one (-0.025, SE = 0.009) coverage plans considering the first 10 years of policy change compared to states without such laws. Looking at the last five years of policy change, we saw increases in effect size and statistical significance. Sensitivity analyses suggest findings are robust to our model specifications.

**Discussion:** Adoption of a medical cannabis law may contribute to decreases in healthcare costs. This phenomenon is likely a secondary effect and suggests positive externalities outside of medical cannabis patients.

**Key Points:**

- States that passed medical cannabis laws, compared to states that did not pass such laws, saw decreases in the average total costs for employer-sponsored health insurance premiums.
- The difference in average total costs for employer-sponsored health insurance premiums, comparing states with and without medical cannabis laws, grew over time.
- This paper contributes to the idea that medical cannabis laws may contain positive externalities.

## 1.0 Introduction

The United States has seen widespread adoption of medical cannabis laws (MCLs) in the past decade.^1,2^ The presence of 38 states with MCLs and 24 with recreational cannabis laws underscores the growing acceptance and utilization of cannabis for medicinal and non-medicinal purposes.^3^ In 2023, the country had approximately 4 million registered medical cannabis patients, with a 4.5 fold increase of 678,408 patients in 2016 to 2,974,433 patients in 2020 in 26 states and Washington, DC.^1,2,4^ This figure is an estimate compiled from state-administered medical cannabis programs and is likely lower than the actual count due to several states not mandating compulsory registration for medical cannabis use and those using cannabis for medical purposes without a registration.

To be eligible for medical cannabis, the majority of states with MCLs maintain a list of qualifying medical conditions.^1^ Prospective patients must secure certification from a physician confirming their diagnosis with one of these conditions to become eligible for medical cannabis use. The implementation of medical cannabis programs in 38 states has significantly increased cannabis usage, necessitating patient registration in most states.

The literature examining the impact of medical cannabis on health conditions suggests cannabis-based medical products likely have efficacy for some conditions and may not be indicated for others.^5–18^ The initial exploration of the effectiveness of medical cannabis was the 2017 national academies of science report on the Health Effects of Cannabis and Cannabinoids.^19^ This report found conclusive evidence that cannabis was effective in treating chronic pain, chemo-therapy induced nausea and vomiting, and multiple sclerosis. It also found some evidence, with newer evidence strengthening their findings of effectiveness for improving short-term sleep outcomes.^6,20–22^ Newer evidence has found strong evidence of efficacy in treating post-traumatic stress disorder,^23–26^ muscle spasticity,^27,28^ and treatment-resistant epilepsy.^13,29,30^

However, medical cannabis may not be medical appropriate for everyone as there may be negative drug-to-drug interaction.^31,32^ For patients with cardiovascular health problems, the inhalation of cannabis smoke can lead to a 20-100% increase in heart rate and an increase in blood pressure and can be contraindicated with blood thiners.^15–18^ Additionally, medical cannabis is likely not indicated for those with schizophrenia.^33^

### 1.1 Medical cannabis laws and Healthcare Costs

Importantly, prior evidence suggests medical cannabis laws may reduce the cost of health care. A study conducted by Cook and Colleagues (2023) estimated the impact of medical cannabis laws on average premium costs in the individual health insurance market from 2010 and 2021. They found that, seven years after adoption, MCLs result in lower health insurer premiums, with a reduction of $-1662.7 after year seven, -$1541.8 after year eight, and $-1625.8 after year nine.^34^

Research has shown that Medicare and Medicaid beneficiaries have fewer prescriptions after a state adopts a medical cannabis law and those fewer prescriptions translate into cost savings.^35,36^ Specifically, research finds a 6% reduction for Medicaid enrollees.^37^ Other evidence of this can be found in the prescription opioid space, where medicinal cannabis laws have reduced the overall prescription of opioids.^38^ Benzodiazepines and antidepressants are other commonly substituted medications. This substitution effect can expand outwards from prescription medications to other mental health/substance-use treatments, which can greatly increase insurance premiums.^39^

There is mixed evidence around the impact of medical cannabis laws on hospitalization rates. Shi (2017) used state-level administrative records from 1997 to 2014, examining the rate of hospitalization involving per 1,000 discharges. Research found medical cannabis laws were associated with 23% reductions in hospitalizations related to opioid dependence or abuse and 13% reduction in opioid pain reliever overdose. They also noted that medical cannabis laws did not increase cannabis-related hospitalizations.^40^ Recent research provides a nuanced examination of the impact of cannabis policy on cannabis-related hospitalizations in Canada. Myran and colleagues (2023) examined cannabis hospitalizations rates per 100,000 population from 2015 to 2021. In the period after Canada’s legalization in 2018 until the COVID-19 pandemic, researchers found significant declines in the rate of cannabis-related hospitalizations. They also found significant increases in cannabis-related hospitalization rates coinciding with the pandemic.^41^

From a causal mechanism perspective, there are likely several reasons why medical cannabis may reduce healthcare costs. Evidence suggests medical cannabis may substitute for traditional prescriptions. Since medical cannabis is not covered by health insurance, the insurer is no longer responsible for paying for the prior prescription nor for its replacement.^42^ Reductions in prescriptions, particularly opioids, may lead to decreased premium costs.

Medical cannabis may also lead to behavioral changes. Cannabis use has been seen to influence various lifestyle factors affecting health, including body-mass index (BMI) and exercise habits. One study involving 20,745 young adult Americans found that cannabis users exercised more frequently than non-users.^43^ Additionally, evidence suggests that cannabis users have lower BMI, potentially aiding in the management of metabolic disorders like obesity and diabetes, which impose significant costs on the US healthcare system.^44^

There is emerging evidence to suggest starting medical cannabis or adopting a medical cannabis law leads to lower alcohol consumption and sales.^45–47^ Yet, this reduction is so far only markedly seen in medical cannabis users specifically, and there are mixed results outside of the context of medicinal use. However, alcohol use is linked to many costly illnesses such as liver failure.

### 1.2 Current Contribution

The current study builds on the existing literature examining the impact of medical cannabis laws on the cost of health insurance. Here, we add the first examination of the impact of medical cannabis laws on the cost of private health insurance plans. To achieve this objective, we used a novel difference-in-difference event-study approach to estimate the cohort-specific average treatment effect on the treated related to medical cannabis law adoption. This method measured the causal impact of medical cannabis law adoption on private health insurance costs overall and over-time, employing both a simple and event-study difference-in-difference approach. We show that states that adopted a medical cannabis law experienced lower average total premium costs per employee for single and employee plus one coverage plans overall and in the last five years of implementation compared to states that did not adopt such laws.

## 2 Methods

We used a quasi-experimental design to estimate the adoption of medical cannabis laws on total average premium costs per employee and total average deductible costs per employee from 2003 to 2022. We estimated the cohort-specific average treatment effect on the treated (C-ATT) related to medical cannabis law adoption using the robust Sun and Abraham (2021) difference-in-difference technique.^48^ We estimated the global C-ATT related to medical cannabis adoption for each outcome separately using a simple DiD and event-study DiD, capturing the overall effect and the dynamic effect of the policy change.

### 2.1 Data sources and Outcomes

This study examined five outcomes, specifically total average premium costs per employee for 1) single, 2) family, and 3) employee plus one health insurance plans as well as total average deductible costs for 4) single and 5) family health insurance plans. All outcome data was ascertained from the Medical Expenditure Panel Survey (MEPS) insurance Component (IC).^49^ The MEPS-IC is a survey of private employers and state and local governments administered annually conducted by Agency for Healthcare Research and Quality. The survey produces state and year indexed estimates of employer-sponsored insurance costs including information on health insurance plan costs. MEPS-IC is publicly available to download. We specifically ascertained state and year indexed estimates of average total premium per enrolled employee at establishments offering health insurance by plan types. Plan types included single, family, and employee plus one. We also ascertained state and year indexed estimates of the average single deductible and the average family deductible. Data on the average employee plus one deductible was not available through MEPS-IC. We log transformed the outcomes to reduce skewness.

We wanted to control for two aspects that might confound the relationship between medical cannabis law adoption and healthcare costs. First, we controlled for population-level demographics potentially related to changes in healthcare costs including the percent of the population male, young, with a high school education or more, white, non-Hispanic, single, living below the poverty line, employed, identified as a veteran, living in in poor health, and with private health insurance. We ascertained these variables from the Current Population Survey through IPUMS.^50^ To control for substance use as a mechanism for changes to healthcare costs, we also included the rate of ethanol consumption and the percent population that reported using cannabis in the past year through the National Institutes of Alcohol Abuse and Alcoholism^51^ and the Substance Abuse and Mental Health Services administration respectively.^52^

Second, to control for the policy environment, we include three dichotomous policy variables for states that expanded Medicaid, states that established their own health insurance marketplace exchanges, and states that later adopted recreational, or adult use, cannabis laws. We ascertained Medicaid expansion dates from the Kaiser Family Foundation.^53^ We ascertained dates of marketplace exchange implementations by triangulating data from healthcare.gov and data from Terrizzi, Mathews-Schults, and Deegan (2022).^54^ We obtained the year of cannabis legalization from the National Conference of State Legistures.^3^ All policies were coded ‘0’ for when a policy was not implemented; states that passed a policy, were coded ‘1’ in the first year of full implementation. We chose these policies as prior evidence suggests individual marketplaces and Medicaid expansion may lower the cost of healthcare. We controlled for states that legalized cannabis so as to not confound the relationship between medical cannabis laws and healthcare costs.

### 2.2 Treatment and Control Units

Our models consist of 22 treatment states (AZ, AR, CT, DE, DC, FL, IL, LA, MD, MN, MO, NH, NJ, NY, ND, OH, OK, PA, RI, UT, VT, and WV) and the District of Columbia and 12 control states (GA, ID, IN, IA, KS, NE, NC, SC, TN, TX, WI, and WY) using 19 time points from 2003-2022. Notably, MEPS-IC data was not collected in 2007. To deal with this, we excluded any covariate data from 2007, skipping the year in the time-series. We set time as an integer starting where 1 = 2003, through 19 = 2022.

We followed the ‘effective date’ of implementation rule for coding changes to medical cannabis laws established by Cook, Sirmans, & Stype (2023) and Anderson & Rees (2023),^34,55^ wherein the medical cannabis law is considered effective when medical sale is operationalized rather than when the law is passed. Notably, there can be large periods of time between when a medical cannabis law is adopted to actual sale. For example, Vermont passed their medical cannabis legislation in 2004.^56^ However, the first dispensary for public sale opened 9 years later in 2013.^57^ Using the effective date of implementation ensures we are properly attributing the impact of the policy on our outcomes. We individually ascertained medical cannabis law adoption dates and law effective dates, seen in Supplemental Table 1.

As our dataset begins in 2003, we included treatment states that had an effective date of 2010 or later. We used this cut point to ensure there were at least 7 years of pre-treatment time to best establish pre-treatment trends between the treated and control cohorts.

As difference-in-difference models require time-variant treatment variables, we excluded states that implemented a medical cannabis law prior to 2003 which included AK, CA, CO, HI, ME, MA, MI, MT, NV, NM OR, and WA. We also excluded states that had effective medical cannabis laws in the year 2022 or had passed a medical cannabis law and had yet to effectively implement it yet which included AL, KY, MS, SD, VA.

### 2.3 Data analysis

Estimating the causal impact of policy is challenging and recent evidence identifies that standard two-way fixed effects models are no longer suitable for identifying the treatment effects of policy change that occurs in multiple time periods.^58^ These models introduce bias when calculating the average treatment effect on the treated (ATT) in a staggered policy adoption setting, such as we have for this analysis. We elected to use a robust Difference-in-Difference (DiD) estimator created by Sun and Abraham (2020) to understand the impact of medical cannabis laws on healthcare costs.

Event-study DiD holds the following assumptions necessary for causal inference: 1) parallel trends in baseline outcomes prior to treatment, 2) no anticipation of treatment, 3) homogenous treatment across time. Prior evidence suggests there may be heterogeneous treatment effects related to medical cannabis laws, likely related to when laws were passed. Sun and Abraham (SA) DiD is an interaction weighted estimator that constructs confidence intervals (CI) of the estimation of treatment effects assuming staggered policy adoption and is robust to heterogeneous treatment effects. The SA DiD can be thought of as a cohort-specific ATT (C-ATT), wherein the average treatment effect on the treated is calculated by weighting the individual cohort’s ATT based on length of panel and cohort size. Otherwise said, the C-ATT can be thought of as a group-time average treatment effect (see Equation 3 in Sun and Abraham 2021).

For this study, we investigated the simple DiD and the dynamic, or event-study, DiD. We estimated the simple DiD by taking a linear combination of the post-treatment lags using the `lincom` post-estimation command in Stata. We did this using the 10 post-treatment lag periods (Overall DiD) as well as the last five post-treatment lag periods (Last Five Years DiD). We present event-study DiD estimates per year for -10 years prior and +10 years post-effective implementation of medical cannabis laws. We test for the assumption of the parallel line trend using the test post-estimation command in Stata using the 10 pre-implementation leads. We provide the cohort-specific weights used for the calculation of the C-ATT and average them over the non-zero time periods to investigate how the C-ATT was calculated. Using the Overall DiD and the Last Five Years DiD effect estimates, we translated the effect on the log scale into real dollars.

We elected to use never treated states, or states that never adopted medical cannabis. States that never adopted medical cannabis likely serve as a superior counterfactual from which to draw inference compared to states that adopted medical cannabis in the last year of the study period. In our analysis, a not yet treated group defined as states with effective medical cannabis laws as of 2021, would consist of only n = 1 state, which is likely not a strong enough comparison group from which to draw inference. We provide trend lines of our five outcomes from 2003 to 2022.

#### 2.3.1 Robustness checks

We used a random treatment assignment for treatment states as a falsification test similar to Cook et. al., (2023). Here we randomly selected a treatment time, drawing a number between 7 and 18 using a random number generator (with 7 equaling the year 2010 and 18 equaling the year 2021) for each treatment state. In this in-time placebo test, we would not expect not to observe an association between medical cannabis laws and healthcare costs. All analyses were conducted in Stata version 18.0 using the `eventstudyinteract` command.^59^

## 3.0 Results

Figure 1 provides overall trend lines for our five health insurance cost outcomes. We see that all aspects of average total health insurance costs increased over the period of time between 2003 to 2022 though at varying rates. The cost of total average premiums for single (111%), family (139%), and employee plus one plans (126%) and the cost of deductible plans for single (304%) and family (261%) all increased from 2003 to 2022.

**Figure 1:**
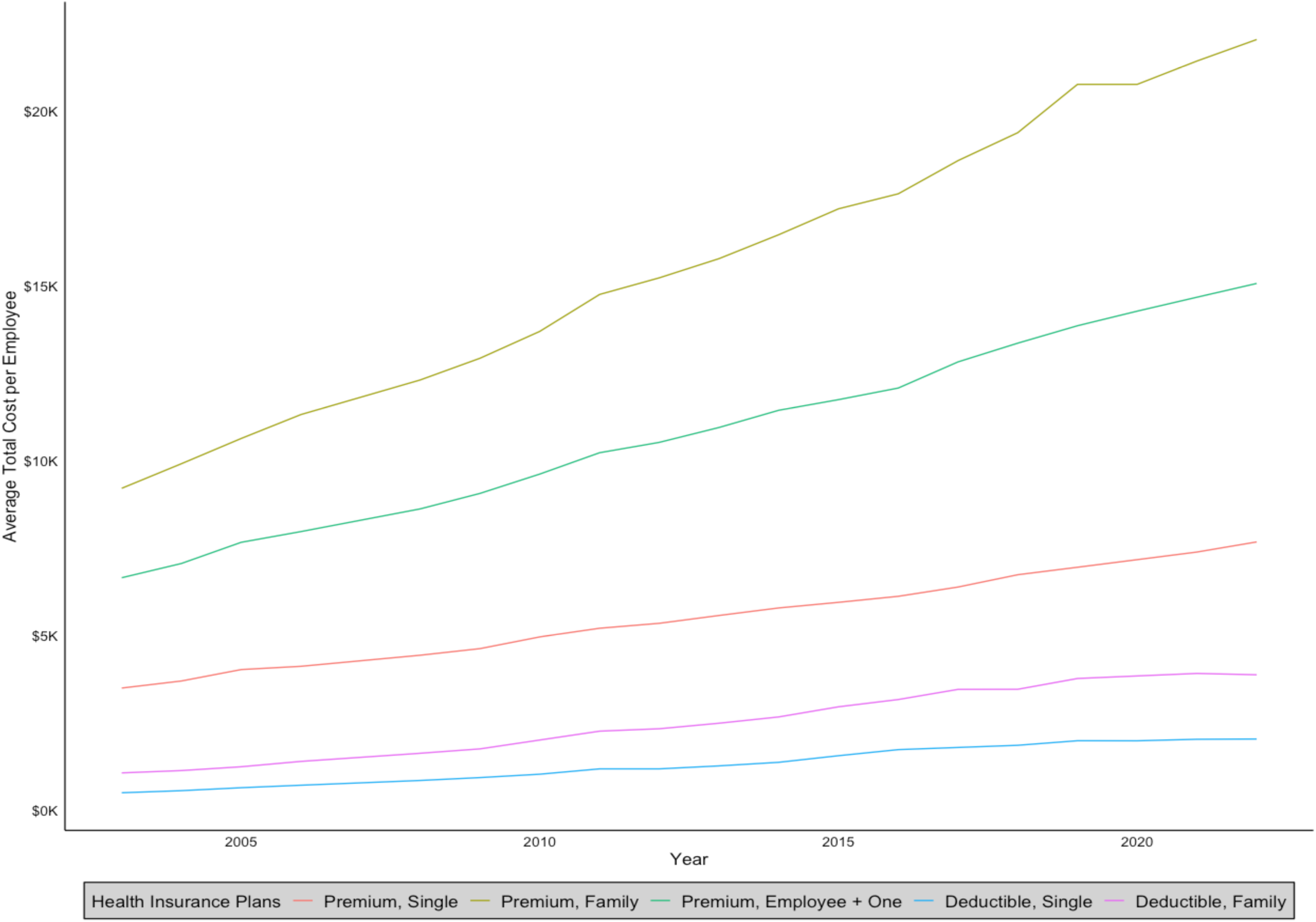
Trends of average total cost per employee for health insurance premiums (single, family, and employee plus one coverage plans) and average total cost per employee for deductibles (single and family coverage plans).

Results of the overall DiD analysis suggest the adoption of medical cannabis laws likely decreased the cost of health insurance for single coverage plans and employee plus one coverage plans (Table 1). Looking at the Overall C-ATT and SE, we note that the adoption of medical cannabis laws was statistically significantly associated with a 3.4% decrease in the average total premium cost for single coverage plans (C-ATT = -0.034, Standard error (SE) = 0.009) and an 2.5% decrease in the average total premium cost for employee plus one coverage plans (ATT = - 0.025, SE = 0.009). Other outcomes did not see statistically significant relations associated with medical cannabis law adoption.

**Table 1:** The impact of medical cannabis law adoption on total premium and deductible costs, 2003-2022. Simple difference-in-difference of cohort-specific average treatment effect on the treated for full models and placebo models.

|  | Full Model |  |  |  | Placebo Model |  |  |  |
| --- | --- | --- | --- | --- | --- | --- | --- | --- |
|  | Overall<br>(n = 22) |  | Last Five Years<br>(n = 13) |  | Overall<br>(n = 22) |  | Last Five Years<br>(n = 13) |  |
|  | CATT | SE | CATT | SE | CATT | SE | CATT | SE |
| *Total Premium Costs* |  |  |  |  |  |  |  |  |
| Single Plan | -0.034* | 0.009 | -0.061* | 0.014 | 0.005 | 0.009 | 0.005 | 0.014 |
| Family Plan | -0.014 | 0.014 | -0.029 | 0.020 | 0.001 | 0.012 | 0.002 | 0.016 |
| Employee Plus<br>One Plan | -0.025* | 0.009 | -0.037* | 0.015 | -0.001 | 0.007 | -0.002 | 0.011 |
| *Deductible* |  |  |  |  |  |  |  |  |
| Single Plan | -0.039 | 0.023 | -0.061 | 0.036 | 0.029 | 0.022 | 0.046 | 0.029 |
| Family Plan | -0.038 | 0.023 | -0.037 | 0.035 | 0.036 | 0.029 | 0.041 | 0.038 |

**Note:** \* p<0.05. Standard errors clustered at the state level with state and year fixed effects included. C-ATT presented here are the result of the `lincom` command in Stata. Overall is the average of all post-treatment years (1 through 10); Last Five Years is the average of years 6 through 10 post-medical cannabis implementation. Never treated as the comparison cohort. Models weighted by population and included percent of the population male, young, with a high school education, white non-Hispanic, single, living below the poverty line, employed, veteran, in poor health, with private health insurance, and reported past year cannabis use. Models also control for ethanol consumption rate with binary policy variables representing the presence of an individual state health insurance marketplace, expansion of Medicaid, and legalization of cannabis for adult use. Placebo Models use in-time random treatment assignment as a falsification test.

Looking at the C-ATT for the last five years of implementation, we see stronger results both in directionality and significance. The average total premium costs for single coverage plans decreased 6.1% (C-ATT = -0.061, SE = 0.014) and decreased 3.7% (C-ATT = -0.037, SE 0.015) for employee plus one coverage plans, both statistically significant. Results suggest that medical cannabis law adoption was not associated changes in average total premium costs for family coverage plans as well as deductibles for single and family plans in the last five years of adoption.

Figure 2 provides the event study plots related to the simple DiD results. For this plot, ‘0’ represents the year the law was effectively implemented with ‘1’ representing the first full year of implementation. Within Figure 2, Plot A is average total premium costs for a single plan; Plot B is average total premium costs for a family plan; Plot C is average total premium costs for an employee plus one plan; Plot D is average total deductible costs for single plans; Plot E is average total deductible costs for family plans. The strong relationship seen between single plans is visible in Plots A and D. For these plots, and to some extent Plot B, we note a somewhat muted effect in the first several years after effective implementation, proceeding by a decrease in effect estimates over time. We provide the post-treatment yearly C-ATT and SE for each outcome in Supplemental Table 2.

**Figure 2:**
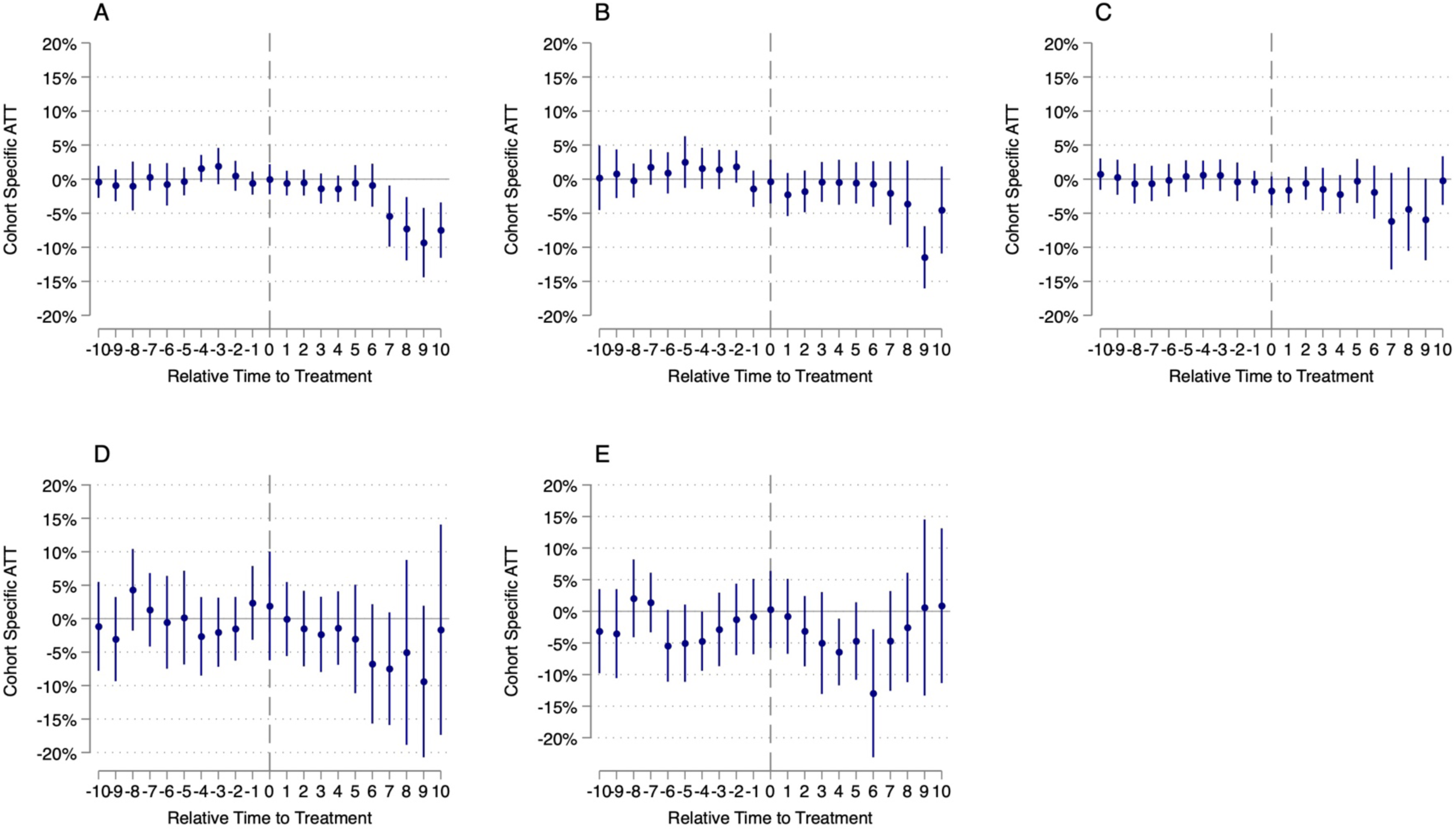
The impact of medical cannabis law adoption on total premium and deductible costs, 2003-2022. Event-study difference-in-difference of cohort-specific average treatment effect on the treated for full models. Plot A is average premium costs for a single plan; Plot B is average premium costs for a family plan; Plot C is average premium costs for an employee plus one plan; Plot D is average deductible costs for single plans; Plot E is average deductible costs for family plans.

Supplemental Table 3 provides the yearly C-ATT and SE for each outcome in the pre-treatment time period and p-values testing the parallel trend assumption. Notably, visible inspection of the trend lines in Figure 2 suggest all of the outcomes appear not to violate the parallel trend assumption. Said otherwise, the 95% confidence intervals include 0 in most pre-treatment periods. Results of the χ2 test of difference from 0 suggest this to be true as all p-values are above p<0.05 (Supplemental Table 3).

Supplemental Table 4 provides the cohort-specific weights used in each of the C-ATT estimation. We provide the weights overtime and as an overall average. We note that the cohort with the largest number of states (2016: AR, FL, NH, and NY) provides the largest weight overall. As expected in event studies, the proportion of cohorts that contribute to the event study effect is related to how long a policy had been in place. Notably, a large portion of states contributed at least some data, specifically cohorts 2011 (AZ = 5 years), 2012 (NJ = 5 years), 2013 (DC & MA = 4 years), 2014 (CT = 3 years), 2015 (DE, IL, & MN = 2 years), and 2016 (AR, FL, NH, and NY = 1 year) to the effect estimates in the last five years.

In Supplemental Table 5, we translated these effects found in Table 1 from the C-ATT on the log scale into actual dollars. For single coverage plans, the total premium costs per employee declined -$238 (95% Confidence Interval (CI) = $234, $242) overall and declined -$460 in the last five years (95% CI = -$450, -$471). For employee plus one coverage plans, the total premium costs per employee declined -$348 (95% Confidence Interval (CI) = $342, $354) overall and declined -$557 in the last five years (95% CI = -$545, -$570). Said otherwise, a company with 50 employees in a state with medical cannabis, assuming a 1:1 ratio of single to employee plus one health insurance plans (ignoring family plans), could have expected to save $14,650 per year over the past 10 years compared to a company in a state without medical cannabis.

We conducted a random-treatment assignment placebo test to examine our models’ robustness. We provide the C-ATT and SE overall and in the last five years after effective implementation of medical cannabis laws in Table 2. We also provided the event-study trend plots for the placebo treatment in Figure 3. Placebo models did not display any statistically significant relationship between the effective implementation of medical cannabis laws and our health insurance outcomes.

**Figure 3:**
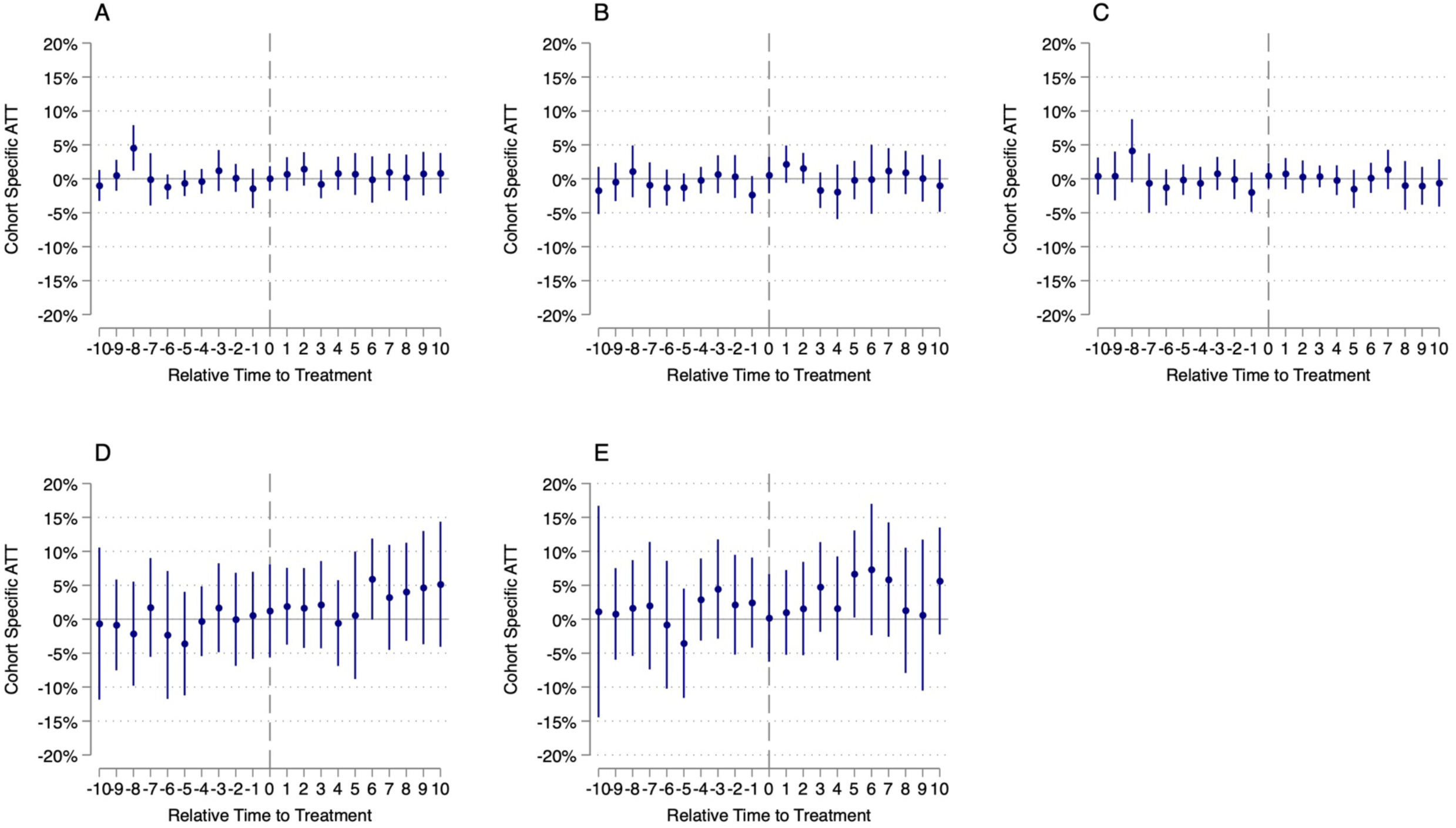
The impact of medical cannabis law adoption on total premium and deductible costs, 2003-2022. Event-study difference-in-difference of cohort-specific average treatment effect on the treated for placebo models. Plot A is average total premium costs for a single plan; Plot B is average total premium costs for a family plan; Plot C is average premium costs for an employee plus one plan; Plot D is average deductible costs for single plans; Plot E is average deductible costs for family plans.

## 4.0 Discussion

In this study we estimated the impact of adopting a medical cannabis law on private health insurance costs using average total premium costs per employee and average total deductible costs per employee as approximate measures. We found evidence of statistically significant reductions over the first 10 years of policy adoption in the average total premium per single and employee plus one coverage plans. Additionally, we find statistically significant reductions for these outcomes in the sixth through tenth years after the implementation of medical cannabis laws. No other outcomes displayed statistically significant increases in the cost of private health insurance. Because of the pooled nature of insurance, lower average total premiums benefit both medical cannabis patients as well as non-patients in states where medical cannabis is legal. Our results are significant as healthcare costs, primarily driven by the cost of premiums, have grown in the past decade plus and account for an increasing proportion of an employer and employees’ budget.

We sought to understand the financial ramifications of our findings on healthcare expenditure GDP under a hypothetical scenario; what would have happened had all 50 states adopted medical cannabis at the same time. According to the US Census Bureau, 304 million people had health insurance in 2022 with 54.5% having a private, employer-sponsored plan (165 million).^60^ Using data from the AHRQ, we assumed that 48.9% of those employer-sponsored plans are single plans (80.6 million) and 18.0% are employee plus one coverage plans (29.7 million).^61^ We assumed within both single coverage plans and employee plus one coverage plans the employer contribution is, on average, around 78%.^62^ Using savings estimates from Supplemental Table 5, we estimated the savings for employers and employees. Had all 50 states implemented medical cannabis at the same time, employers may have experienced a total savings of $14.9 billion for single coverage plans and $8 billion for employee plus one coverage plans in a given year; Employees may have seen a total savings of $4.2 billion for single plans and $2.3 billion for employee plus one plans in a given year. Under this assumption, medical cannabis laws could have reduced healthcare expenditure GPD by 0.56% in 2022 had all states adopted this policy change at the same time.^63^

Our findings align with previous research suggesting that medical cannabis laws may reduce health insurance premiums in individual health insurance markets.^34^ Here, we expand upon this literature and find that medical cannabis laws also likely decrease the cost of private, employer-sponsored health insurance for both single and employee plus one coverage plans. Our findings are consistent with prior research that found medical cannabis laws were associated with lower prescription opioid prescribing.^35–38,64^ Importantly, prior research indicates that the effect of medical cannabis laws likely took several years to manifest.^34^ This is a logical finding as behavior change associated with public policy typically requires time, especially when removing a restrictive policies, such as legalizing the medical sale of cannabis. Additionally, since we are estimating the impact of these laws on secondary outcomes—health insurance costs associated with the health benefits of medical cannabis use—it is understandable that the full impact may not be felt for several years after the policies take effect.

Additionally, this study builds on the work of Anderson and Rees (2023) and Cook et al., (2023) in developing a standard for evaluating cannabis policy.^34,55^ For each state, we provide the year of law enactment and the year the policy was effective (Supplemental Table 1). We agree with prior research that suggests using the year a medical cannabis law was enacted to evaluate its impact likely leads to misspecified models as there is typically time between policy enactment and availability of cannabis products.

It appears our analyses are robust to DiD model assumptions. First, based on tests of pre-treatment parallel trend differences suggest no difference between treatment and control for our outcomes (Supplemental Table 3). Second, examination of event-study trend lines does not show large departures prior to treatment periods implying there was likely no anticipation of treatment. Third, the Sun and Abraham (2021) method is protective against treatment heterogeneity. Moreover, the results of our placebo testing suggest our models are correctly specified for our outcomes and policy change.

### 4.1 Limitations

There are limitations to our study. As our study period began in 2003, we were unable to include states with time-invariant medical cannabis laws which excluded early policy adopters. Difference-in-Difference analyses require pre-trend time periods to capture policy counterfactual. Thus, we set an a priori threshold of having 7 years of pre-treatment. This further excluded states that had an effective medical cannabis law after 2003 but before 2010. Therefore, our results here are specific to states that had effective medical cannabis laws after 2010.

Selection bias affects the majority of state-level policy analyses. Here, our findings are impacted by the states that did and did not choose to adopt medical cannabis laws. Additionally, our analyses are biased by potential unmeasured confounding. Our models included covariates that likely confound the relationship between medical cannabis laws and health care costs including the percent of the population that reported past year prior cannabis use, the presence/absence of Medicaid expansion, state-run individual health insurance marketplaces, and recreational cannabis laws among others. It is possible some other unmeasured confounder is biasing our results.

### 4.2 Conclusion

We evaluated the impact of medical cannabis law adoption on employer-sponsored health insurance costs through average total premium and average total deductible costs per employee from 2003-2022. To this end, we utilized a heterogeneity robust Difference-in-Difference approach. Our findings suggest that medical cannabis laws likely decreased the cost of both single and employee plus one coverage plans for average total premium costs. Moreover, the impact of these laws on private health insurance costs is evident five years after the law becomes effective. States should consider these positive externalities associated with medical cannabis legalization when considering whether or not to adopt a medical cannabis law.

## Data Availability

The outcome data used for this study is publicly available from the Agency for Healthcare Research and Quality.

https://datatools.ahrq.gov/meps-ic/?type=tab&tab=mepsich3ps&_gl=1%2A1jfniqe%2A_ga%2AMTk3NDY5MzY5MS4xNzAxNzg1MzE0%2A_ga_1NPT56LE7J%2AMTcwMTg2NzY5Mi4yLjAuMTcwMTg2NzY5Mi4wLjAuMA.

## Acknowledgements

No external funding was used in the creation of this work.

## Data Availability Statement

The outcome data used for this study is publicly available from the Agency for Healthcare Research and Quality and can be found here: https://datatools.ahrq.gov/meps-ic/?type=tab&tab=mepsich3ps&_gl=1%2A1jfniqe%2A_ga%2AMTk3NDY5MzY5MS4xNzAxNzg1MzE0%2A_ga_1NPT56LE7J%2AMTcwMTg2NzY5Mi4yLjAuMTcwMTg2NzY5Mi4wLjAuMA.

