## Supplemental Material for "Measuring the Impact of Medical Cannabis Law Adoption on Employer-sponsored Health Insurance Costs: A Difference-in-Difference analysis, 2003-2022"

**Supplemental Table 1:** Medical cannabis law adoption and effectiveness years

| State | Year Date of Medical Law Passed[1] | Year Medical Law was Effective[2] |
|---|---|---|
| Alabama^ | 2021 | Not yet implemented |
| Alaska* | 1998 | 1998 |
| Arizona | 2010 | 2011 |
| Arkansas | 2016 | 2016 |
| California* | 1996 | 1996 |
| Colorado* | 2000 | 2000 |
| Connecticut | 2012 | 2014 |
| Delaware | 2011 | 2015 |
| District of Columbia | 1998 | 2013 |
| Florida | 2016 | 2016 |
| Georgia | --- | --- |
| Hawaii* | 2000 | 2000 |
| Idaho | --- | --- |
| Illinois | 2013 | 2015 |
| Indiana | --- | --- |
| Iowa | --- | --- |
| Kansas | --- | --- |
| Kentucky^ | 2023 | Not yet implemented |
| Louisiana | 2017 | 2019 |
| Maine* | 1999 | 1999 |
| Maryland | 2014 | 2017 |
| Massachusetts | 2012 | 2013 |
| Michigan* | 2008 | 2008 |
| Minnesota | 2014 | 2015 |
| Mississippi^ | 2022 | Not yet implemented |
| Missouri | 2018 | 2020 |
| Montana* | 2004 | 2004 |
| Nebraska | --- | --- |
| Nevada* | 2000 | 2001 |

| | | |
|---|---|---|
| New Hampshire | 2013 | 2016 |
| New Jersey | 2010 | 2012 |
| New Mexico* | 2007 | 2007 |
| New York | 2014 | 2016 |
| North Carolina | --- | --- |
| North Dakota | 2016 | 2019 |
| Ohio | 2016 | 2019 |
| Oklahoma | 2018 | 2018 |
| Oregon* | 1998 | 1998 |
| Pennsylvania | 2016 | 2018 |
| Rhode Island* | 2006 | 2006 |
| South Carolina | --- | --- |
| South Dakota^ | 2020 | Not yet implemented |
| Tennessee | --- | --- |
| Texas | --- | --- |
| Utah | 2018 | 2020 |
| Vermont* | 2004 | 2004 |
| Virginia | 2020 | 2020 |
| Washington* | 1998 | 1998 |
| West Virginia | 2017 | 2017 |
| Wisconsin | --- | --- |
| Wyoming | --- | --- |

**Note:** *Denotes early medical cannabis law adopters; ^denotes late medical cannabis law adopters. Both were not included in the analysis. Law passage years were obtained from the National Conference of State Legislatures (https://www.ncsl.org/health/state-medical-cannabis-laws). Medical cannabis law effective dates were obtained from Anderson & Rees (2023).

**Supplemental Table 2:** Post-treatment event-study cohort-specific average treatment effect on the treated

| | Post-Treatment Year | | | | | | | | | | |
|---|---|---|---|---|---|---|---|---|---|---|---|
| **C-ATT** | 0 | 1 | 2 | 3 | 4 | 5 | 6 | 7 | 8 | 9 | 10 |
| *Average Total Premium* | | | | | | | | | | | |
| Single Plan, | 0.000 | -0.006 | -0.005 | -0.014 | -0.014 | -0.006 | -0.009 | -0.054 | -0.073 | -0.093 | -0.075 |
| SE | 0.011 | 0.009 | 0.010 | 0.011 | 0.010 | 0.013 | 0.016 | 0.023 | 0.024 | 0.026 | 0.021 |
| Family Plan | 0.007 | -0.008 | -0.007 | -0.005 | 0.006 | 0.008 | 0.001 | -0.017 | -0.038 | -0.074 | -0.018 |
| SE | 0.014 | 0.014 | 0.016 | 0.017 | 0.016 | 0.016 | 0.017 | 0.027 | 0.032 | 0.025 | 0.037 |
| Employee + One Plan | -0.017 | -0.016 | -0.006 | -0.015 | -0.022 | -0.003 | -0.019 | -0.062 | -0.044 | -0.059 | -0.002 |
| SE | 0.011 | 0.010 | 0.012 | 0.016 | 0.014 | 0.017 | 0.020 | 0.036 | 0.031 | 0.031 | 0.018 |
| *Average Total Deductible* | | | | | | | | | | | |
| Single Plan | 0.019 | -0.001 | -0.015 | -0.023 | -0.014 | -0.030 | -0.068 | -0.075 | -0.050 | -0.094 | -0.017 |
| SE | 0.042 | 0.028 | 0.029 | 0.029 | 0.028 | 0.041 | 0.046 | 0.043 | 0.071 | 0.058 | 0.080 |
| Family Plan | 0.003 | -0.008 | -0.031 | -0.050 | -0.064 | -0.047 | -0.130 | -0.047 | -0.026 | 0.006 | 0.009 |
| SE | 0.031 | 0.030 | 0.028 | 0.041 | 0.027 | 0.031 | 0.052 | 0.040 | 0.044 | 0.071 | 0.062 |

**Note:** Year = 0 represents the year of medical cannabis law is effective (see Supplemental table 1 for states and dates). Year = 1 represents the first full year of implementation. Overall effect in the first 10 years of implementation calculated using time points 1 through 10.

**Supplemental Table 3:** Pre-treatment event-study cohort-specific average treatment effect on the treated.

| | Pre-treatment Years | | | | | | | | | | |
|---|---|---|---|---|---|---|---|---|---|---|---|
| **C-ATT** | -10 | -9 | -8 | -7 | -6 | -5 | -4 | -3 | -2 | -1 | Pre-treatment Trends, P-value |
| *Average Total Premiums* | | | | | | | | | | | |
| Single Plan | -0.004 | -0.009 | -0.010 | 0.003 | -0.008 | -0.003 | 0.016 | 0.019 | 0.005 | -0.006 | 0.137 |
| SE | 0.012 | 0.012 | 0.018 | 0.010 | 0.016 | 0.011 | 0.010 | 0.014 | 0.011 | 0.009 | |
| Family Plan | 0.013 | 0.005 | -0.002 | 0.016 | 0.024 | 0.022 | 0.024 | 0.022 | 0.018 | -0.006 | 0.530 |
| SE | 0.018 | 0.015 | 0.011 | 0.015 | 0.014 | 0.019 | 0.013 | 0.014 | 0.012 | 0.013 | |
| Employee + One Plan | 0.007 | 0.003 | -0.006 | -0.006 | -0.001 | 0.004 | 0.006 | 0.006 | -0.004 | -0.004 | 0.980 |
| SE | 0.012 | 0.013 | 0.015 | 0.013 | 0.012 | 0.012 | 0.011 | 0.012 | 0.014 | 0.008 | |
| *Average Total Deductible* | | | | | | | | | | | |
| Single Plan | -0.011 | -0.031 | 0.043 | 0.013 | -0.005 | 0.002 | -0.026 | -0.020 | -0.015 | 0.024 | 0.404 |
| SE | 0.034 | 0.032 | 0.031 | 0.028 | 0.035 | 0.036 | 0.030 | 0.026 | 0.024 | 0.028 | |
| Family Plan | -0.031 | -0.035 | 0.020 | 0.014 | -0.055 | -0.050 | -0.047 | -0.029 | -0.013 | -0.008 | 0.069 |
| SE | 0.034 | 0.036 | 0.031 | 0.024 | 0.029 | 0.031 | 0.024 | 0.030 | 0.029 | 0.030 | |

**Note:** Pre-treatment trends test is result of `lincom` in Stata which tests whether the combination of pre-treatment years -10 through -1 is statistically different from 0 using a chi2 test.

**Supplemental Table 4:** Cohort-specific weights for average treatment effect on the treated calculation

| | Cohorts for Effective Implementation of Medical Cannabis Laws | | | | | | | | | |
|---|---|---|---|---|---|---|---|---|---|---|
| **Time** | **2011** | **2012** | **2013** | **2014** | **2015** | **2016** | **2017** | **2018** | **2019** | **2020** |
| -10 | --- | --- | --- | --- | 0.161 | 0.350 | 0.065 | 0.139 | 0.142 | 0.144 |
| -9 | --- | --- | --- | 0.029 | 0.156 | 0.342 | 0.062 | 0.134 | 0.138 | 0.139 |
| -8 | --- | --- | 0.056 | 0.027 | 0.147 | 0.325 | 0.058 | 0.125 | 0.131 | 0.131 |
| -7 | --- | 0.064 | 0.051 | 0.026 | 0.137 | 0.305 | 0.054 | 0.119 | 0.121 | 0.123 |
| -6 | 0.038 | 0.061 | 0.049 | 0.025 | 0.132 | 0.293 | 0.053 | 0.115 | 0.116 | 0.119 |
| -5 | 0.039 | 0.061 | 0.048 | 0.024 | 0.131 | 0.293 | 0.053 | 0.116 | 0.116 | 0.119 |
| -4 | 0.040 | 0.061 | 0.048 | 0.024 | 0.131 | 0.293 | 0.053 | 0.114 | 0.116 | 0.119 |
| -3 | 0.042 | 0.059 | 0.048 | 0.024 | 0.131 | 0.294 | 0.053 | 0.113 | 0.116 | 0.119 |
| -2 | 0.044 | 0.059 | 0.049 | 0.024 | 0.130 | 0.294 | 0.053 | 0.113 | 0.116 | 0.118 |
| -1 | 0.045 | 0.060 | 0.049 | 0.024 | 0.130 | 0.295 | 0.053 | 0.113 | 0.115 | 0.119 |
| 0 | 0.044 | 0.059 | 0.049 | 0.024 | 0.130 | 0.298 | 0.052 | 0.111 | 0.113 | 0.120 |

| | | | | | | | | | | |
|---|---|---|---|---|---|---|---|---|---|---|
| 1 | 0.044 | 0.059 | 0.049 | 0.024 | 0.129 | 0.298 | 0.052 | 0.110 | 0.114 | 0.120 |
| 2 | 0.044 | 0.059 | 0.049 | 0.024 | 0.127 | 0.300 | 0.052 | 0.111 | 0.114 | 0.120 |
| 3 | 0.050 | 0.068 | 0.056 | 0.027 | 0.145 | 0.339 | 0.060 | 0.125 | 0.129 | --- |
| 4 | 0.057 | 0.077 | 0.064 | 0.031 | 0.167 | 0.393 | 0.068 | 0.143 | --- | --- |
| 5 | 0.068 | 0.089 | 0.076 | 0.034 | 0.196 | 0.458 | 0.079 | --- | --- | --- |
| 6 | 0.075 | 0.098 | 0.083 | 0.039 | 0.210 | 0.496 | --- | --- | --- | --- |
| 7 | 0.152 | 0.191 | 0.168 | 0.077 | 0.413 | --- | --- | --- | --- | --- |
| 8 | 0.263 | 0.331 | 0.277 | 0.129 | --- | --- | --- | --- | --- | --- |
| 9 | 0.299 | 0.385 | 0.316 | --- | --- | --- | --- | --- | --- | --- |
| 10 | 0.438 | 0.562 | --- | --- | --- | --- | --- | --- | --- | --- |
| ***Average Weights*** | | | | | | | | | | |
| | 0.105 | 0.133 | 0.088 | 0.035 | 0.161 | 0.333 | 0.058 | 0.120 | 0.121 | 0.124 |

**Note:** 2011 cohort consists of Arizona, 2012 cohort consists of NJ, 2013 consists of DC and MA, 2014 consists of CT, 2015 consists of DE, IL, and MN, 2016 consists of AR, FL, NH, and NY, 2017 consists of MD and WV, 2018 consists of OK and PA, 2019 consists of LA, ND, and OH, 2020 consists of MO, UT, and VA. Average weights = average from non-zero weights. As year passage and time was consistent across all models, the weighting provided here was also consistent.

**Supplemental Table 5**: The impact of medical cannabis law adoption on average total premium and deductible costs per employee translated into dollars, 2003-2022.

**Full Model**

| | Overall (n = 22 States) | | | | | Last Five Years (n = 13) | | | | |
|---|---|---|---|---|---|---|---|---|---|---|
| | *Average $* | *SE* | *Effect* | *95% CI* | | *Average $* | *SE* | *Effect* | *95% CI* | |
| | | | | *Lower* | *Upper* | | | | *Lower* | *Upper* |
| *Total Premium Costs* | | | | | | | | | | |
| Single Plan | $7,000 | $62 | -$238* | -$234 | -$242 | $7,545 | $86 | -$460* | -$450 | -$471 |
| Family Plan | $20,250 | $185 | -$283 | -$278 | -$289 | $22,053 | $223 | -$640 | -$626 | -$653 |
| Employee + One Plan | $13,910 | $122 | -$348* | -$342 | -$354 | $15,059 | $168 | -$557* | -$545 | -$570 |
| *Deductible* | | | | | | | | | | |
| Single Plan | $1,826 | $28 | -$71 | -$69 | -$73 | $1,927 | $45 | -$118 | -$112 | -$123 |
| Family Plan | $3,506 | $53 | -$133 | -$129 | -$137 | $3,697 | $77 | -$137 | -$131 | -$143 |

Note: Overall is the average of all post-treatment years for all medical cannabis law adopters; Last Five Years is the average of years 6 through 10 post-medical cannabis implementation. Never treated as the comparison cohort. Average $ is equal to the states that passed medical cannabis laws in their post-treatment period. We then applied the effect size to find the effect in real dollars for the C-ATT and the 95% CI using the standard errors. * Denotes where effect has a p-value <0.05
